# Finding Tentative Causes for the Reduced Impact of Covid-19 on the Health Systems of Poorer and Developing Nations: An Ecological Study of the Effect of Demographic, Climatological and Health-Related Factors on the Global Spread of Covid-19

**DOI:** 10.1101/2020.05.25.20113092

**Authors:** Shabnam Banerjee, Arunanjan Saha

**Affiliations:** Tata Medical Center; University of Calcutta

## Abstract

**Objective:** The objective of this study is to evaluate the association with different factors empirically found to affect the spread and the severity of Covid-19. Evidently there is less likelihood of having one single and absolute solution to this pandemic. It is pragmatic to look for a multi-pronged and collaborative assembly of probable solutions, which is the higher objective of this study.

**Design:** Ecological study.

**Setting:** Global setting including 45 countries from all six inhabited continents

**Population:** Two (2) or three (3) countries from each geographical region of the continents selected on the basis of population

**Main outcome:** measures correlation factors derived from comparisons between different sets of variables

**Results:** Empirical trends suggested in the existing literature were quantified in a global setting establishing clear trends. Correlation between the proportion of the population affected and median age, prime climate zones, malaria and tuberculosis incidence, BCG coverage and mitigation measures were established.

**Conclusions:** The study findings suggest that demographic and climatological factors, high endemicity of TB and Malaria, and universal BCG programmes may have a cushioning effect in the impact of Covid-19 on health systems of poorer and developing nations. In the light of these findings more emphasis is necessary on the protective effects of BCG and antiviral properties of antimalarial drugs.

**Background:** The coronavirus disease (Covid-19) caused by the severe acute respiratory syndrome coronavirus 2 (SARS-CoV-2) has affected 213 countries as of 20th May 2020, with a total of 5,017,883 total cases reported globally, and has caused 325,624 reported deaths till date (worldometers.info). It began with the report of an outbreak of pneumonia of unknown cause in Wuhan, China on 31^st^ December 2019. This outbreak was declared a Public Health Emergency of International Concern on 30th January 2020. Even as early as 18th February 2020, it was noted that inspite of lower case fatality rates Covid-19 had already resulted in more deaths than SARS and MERS combined.^1^ On the 11th of March the WHO made the assessment that Covid-19 could be classified as a pandemic. A systematic review found the basic reproduction number between 2.0 - 3.0.^2^ According to the WHO, the crude case fatality rate is 3%, with 15% of those affected suffering from severe disease and 5% critical.^3^

As developed countries with highly ranked health systems report high burden of cases straining health system capacities, concerns grow about the impact on resource constrained health systems in developing and underdeveloped nations. A better understanding of the disease epidemiology could help in planning the pandemic response, both in terms of resource allocation and mitigation measures in areas and populations more likely to be affected. In the second week of April, 2020, the IMF also was reported to be keen to know the reason behind the lower impact of Covid-19 in African and Asian countries; this also served as a driving factor behind this study.

## Introduction

This study takes a comprehensive look into the global patterns of incidence of Covid-19 with respect to demographic, climatological and health related determinants and mitigation efforts of various countries.

Trends in Italy and South Korea suggest^4^ a dramatically higher burden of mortality from Covid-19 in countries with older populations. Covid-19 related morbidity and mortality rates^5^ are associated with the median ages of the respective countries. WHO surveillance data^6^ shows a progressively higher proportion of cases belonging to the 20-39 age-group, but case fatality rates are seen to increase with age^3^.

There is some concern that high population densities would catalyse the spread of the disease^7^. However, a study of 8 cities with high spread of the disease noted the lack of significant community spread in expected locations based only on high population proximity and extensive interaction through travel^8^.

Studies conducted in Wuhan^9^, in a group of chinese cities^10^, in Jakarta, indonesia^11^ and iran^12^ found that higher temperature, relative humidity, wind speed, and solar radiation reduced the transmission of Covid-19. Significant spread has been noted^8^ in cities and regions lying between 30 and 50 degree latitude with average temperatures of 5-11 degree celsius and low specific and absolute humidity. Between January - March 2020 incidence had peaked in temperate regions of the northern hemisphere in and were lower in both warmer/wetter and colder/dryer regions^13^. However, a United States-based model^14^ of SARS-CoV-2 transmission found that it could proliferate in any time of the year. Other studies^15^ have commented that virus transmission had no seasonality and spanned all climatic zones. A standing committee^16^ of the National Academies of Sciences, Engineering, and Medicine, the USA, stated that factors such as temperature and humidity would not lead to any significant reduction in the disease spread due to the lack of pre-existing immunity globally.

A comparison of distribution maps^17^ of Beta-CoVs diseases (MERS, SARS and COVID-19) and malaria incidence unveiled an inverse relationship. However the WHO^18^ refused to give any reassurance as it reported that the African region which carries 90% of the global Malaria burden had reported 25000 cases of Covid-19 as of 30th April 2020.

The antimalarial agents chloroquine and hydroxychloroquine^19^ have proven in-vitro efficacy in reducing viral replication by increasing endosomial ph and interfering with the glycosylation of the cellular receptor. Guidelines on their use in managing Covid-19 have been released in Guangdong province, Netherlands and Italy^19^. A Cochrane review related to the use of chloroquine or hydroxychloroquine for the prevention and treatment of COVID-19 is also being undertaken^20^. However the WHO^25^ still maintains that there is insufficient data to assess the efficacy of either of chloroquine or hydroxychloroquine in treating or preventing COVID-19^25^.

Universal BCG vaccination^21^ has been associated with reduced morbidity and mortality from COVID 19. However, in a scientific brief published in April 2020 the WHO ^22^ states that there is no evidence confirming that the BCG vaccine protects people against infection with the COVID-19 virus.

There is a large body of evidence that BCG has a nonspecific or heterologous protective effect in addition to TB, especially reducing rates of sepsis, respiratory tract infections and diarrhoea, leading to reduced all cause mortality in children and especially infants.^23^. This protective effect was found to be dramatic and rapid, evident as early as 3 days after BCG vaccination, and can persist for at least 5 years, though it may be negated by subsequent non live vaccines.^30^. Multiple studies demonstrate that BCG vaccination alters the cytokine profile in administered subjects, promotes Th1 polarisation of the developing infant immune system, and increases humeral responses to other vaccines including the influenza vaccine. BCG also has a capacity of inducing trained immunity (TRIM) or innate immune memory which is mediated by epigenetic mechanisms, characterised by increased pro-inflammatory responses to unrelated secondary infections^30^. The epigenetic modifications following BCG vaccination have been found to persist very long and are likely due to DNA methylation which are more stable than histone modifications^30^. Ongoing clinical trials in Australia, New Zealand and the USA are evaluating whether BCG vaccination of health workers could protect them from contracting COVID-19, whether it may prevent severe COVID-19 infection among older adults and a study in Germany is testing whether a recombinant vaccine strain derived from BCG can protect either of the two high risk populations^24^.

Mitigation and suppression strategies and social distancing measuresmay sufficiently reduce transmission so that effective reproduction number R is lowered, as demonstrated by China^25^. The WHO strategy update^3^ on 14th April 2020 observes that physical distancing measures and movement restrictions can slow down COVID-19 transmission. Therefore WHO recommends suppressing community transmission through context appropriate IPC measures, population level physical distancing and appropriate and proportionate restrictions on travel.

The available literature has mostly focused on the association of each of these determinants separately with morbidity and mortality from Covid-19, mostly in regional or national contexts and findings are self-contradictory. This study focuses more on the spread of the disease in terms of proportion of the population affected with Covid-19 rather than morbidity and mortality; it considers all these variables simultaneously to give a more complete picture; with a global context using quantitative analysis to prove empirical trends.

## Methods

The objective of this study was to evaluate the association of different factors empirically found to affect the spread and the severity of Covid-19. An ecological study with a mixed methodological approach was undertaken.This approach was appropriate considering the global scale of the pandemic and the large number of variables that could affect its spread. Qualitative methods were used to analyse climate classification based spread of the disease and the impact of lockdown measures on the epidemiological curve. Quantitative methods were used to find the geographical correlation between selected demographic variables, health determinants and the disease incidence and mortality.

### Data Collection

Data sourced from accredited organisations available in the public domain for 45 countries distributed over each different geographical region of six continents were selected to be studied based on their population. The demographic data including population as on 1st July 2019, life expectancy, population density and median age were collected from the worldometers website^26^ which has sourced its data from the World Population Prospects 2019, published by the United Nations Population Division^27^. The prime climate conditions in order of count for each of these countries according to the Koppen-Geiger classification were noted^28^. From the worldometers website^29^ country-wise statistical data about Covid-19, sourced mainly from government data of the respective countries, as updated till 30th April 2020 was collected; this included the country-wise date of first report, total number of reported cases, date of first death, total deaths, total tests, recovered, highest number of cases in a day, date and number of cases on peak day, and date and number of cases on the day with the second highest case count. The case fatality rate (total deaths with respect to total reported cases), specific mortality rate (per million population), average recovery rate, number of days until and from the spike and number of days from the second highest report in a day till the peak day were calculated for each country. Country-wise data on the incidence of Malaria per thousand population were collected from global health observatory, WHo^30^. Data on incidence of tuberculosis by country were obtained from the World Bank website^31^. BCG coverage, and duration of years of BCG administration for those countries with universal coverage was obtained from WHo data^32^. The nature of combat measures adopted in some of the countries, for example whether selective or stringent lockdown measures were imposed, was found from widely circulated and referred media sources. The data collected is represented in a master table in Appendix 1. Abbreviations have been used for country names, as listed in the table, for the ease of diagrammatic representation.

### Data analysis

Geospatial comparison of Koppen-Geiger classification with Covid-19 case distribution from the WHO dashboard was followed by cross-tabulation of Koppen climate classifications most prevalent in the selected countries with the total number of cases and proportion of the population affected. The association between the quantitative demographic and health-related data with the number of Covid-19 cases in each country was examined using graphical representation followed by bivariate analysis and calculation of correlation coefficients. Similar geographical correlations were done of the data with the percentage of the populations affected and the case fatality rate.

## Results

The percentage of Covid-19 cases in the population has a significant positive correlation with the median age of the respective countries (r= 0.595), as shown in Figure 1. The case fatality rate is also positively associated with the median age (r=0.32).

**Fig 1.**
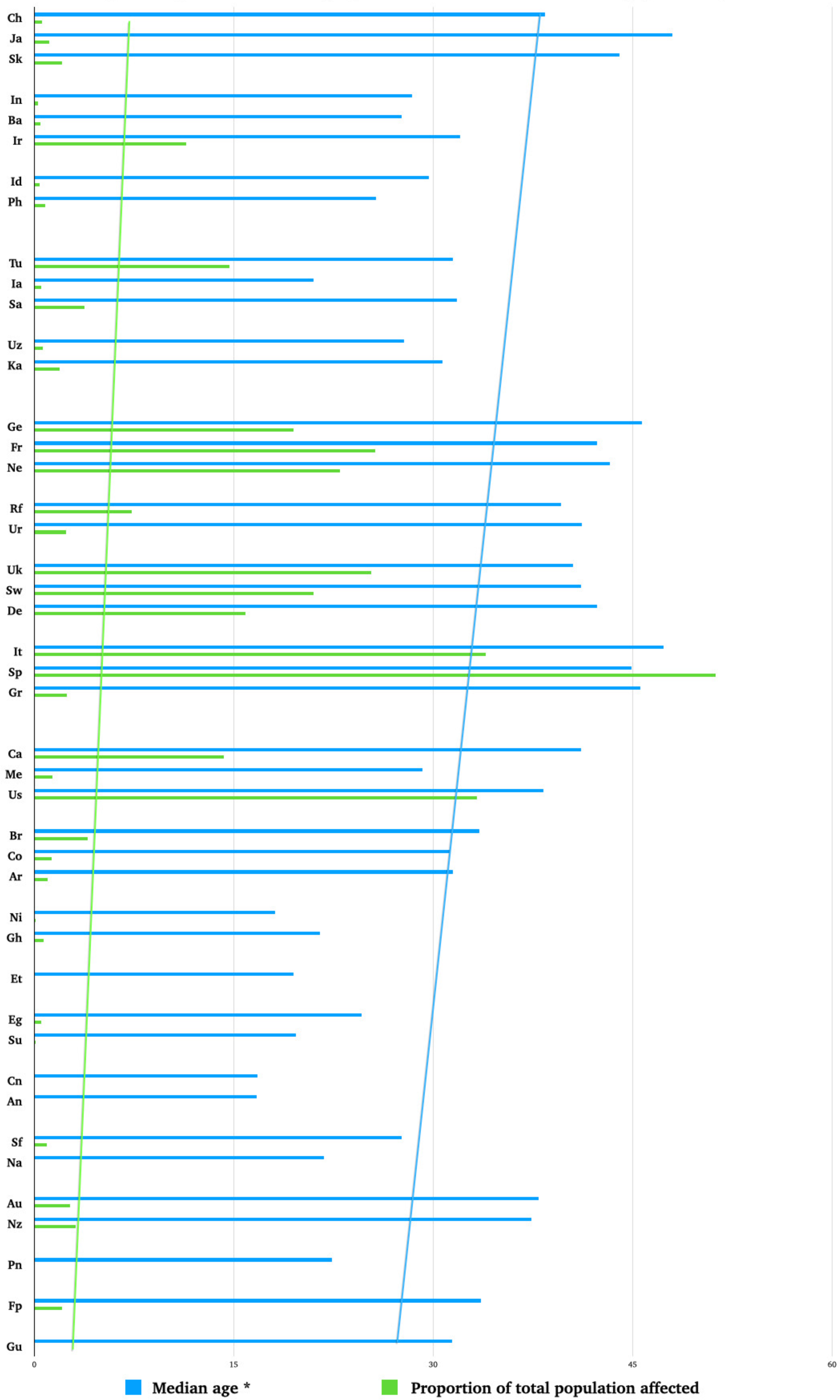
Proportion of total population affected vas Median age, country-wise.

Population density is weakly associated (r=0.16) with the percentage of positive cases among those tested for Covid-19, and has no correlation (r=0) with the total number of cases or with the percentage of the total population affected with Covid-19 (r= - 0.03).

The total number of cases reported varies proportionately with the total number of tests done (r=0.89). However, the percentage of cases among those tested is not significantly associated (r= 0.16) with the total number of tests done.

Cross-tabulation of most prevalent Koppen-Geiger climate conditions with the percentage of the population affected with Covid-19 as shown in Figure 2, reveals that the highest percentage of cases are noted in Oceanic (Cfb), Humid subtropical (Cwa, Cfa), Humid continental (Dfb), Tropical Savannah (Aw), and Semi Arid hot (BSh) climate zones.

**Fig 2:**
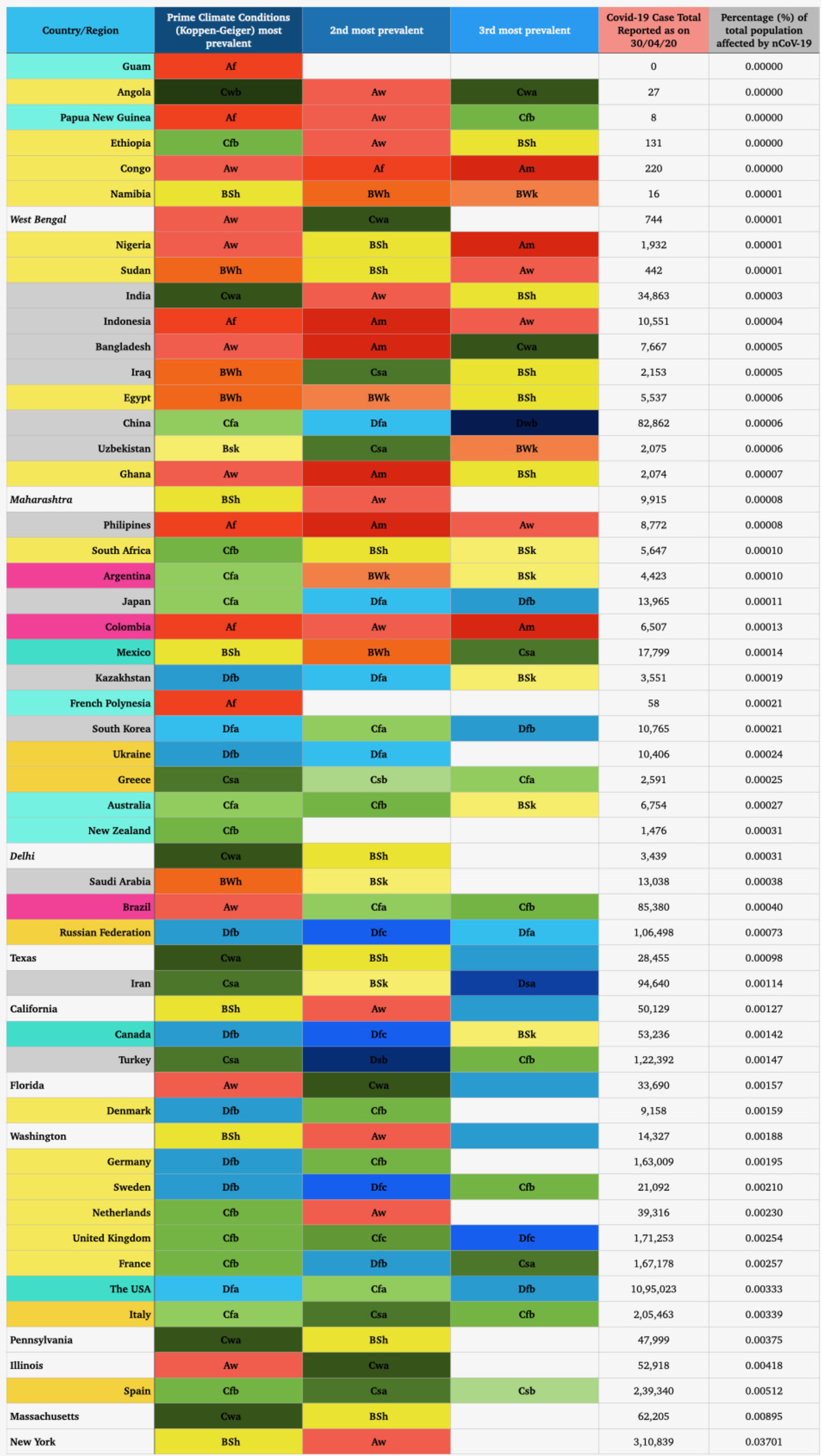
Climate zones most prevalent in Countries by percentage of population affected by nCoV-19.

The percentage of the population affected with Covid-19 is inversely related to the incidence of malaria in that population (r=—0.28), as shown in Figure 3. The total number of reported cases of Covid-19 also decreases slightly with the higher incidence of malaria (r=—0.16). (To calculate these correlation values reported incidence of 0 was substituted as 0.001).

**Fig 3.**
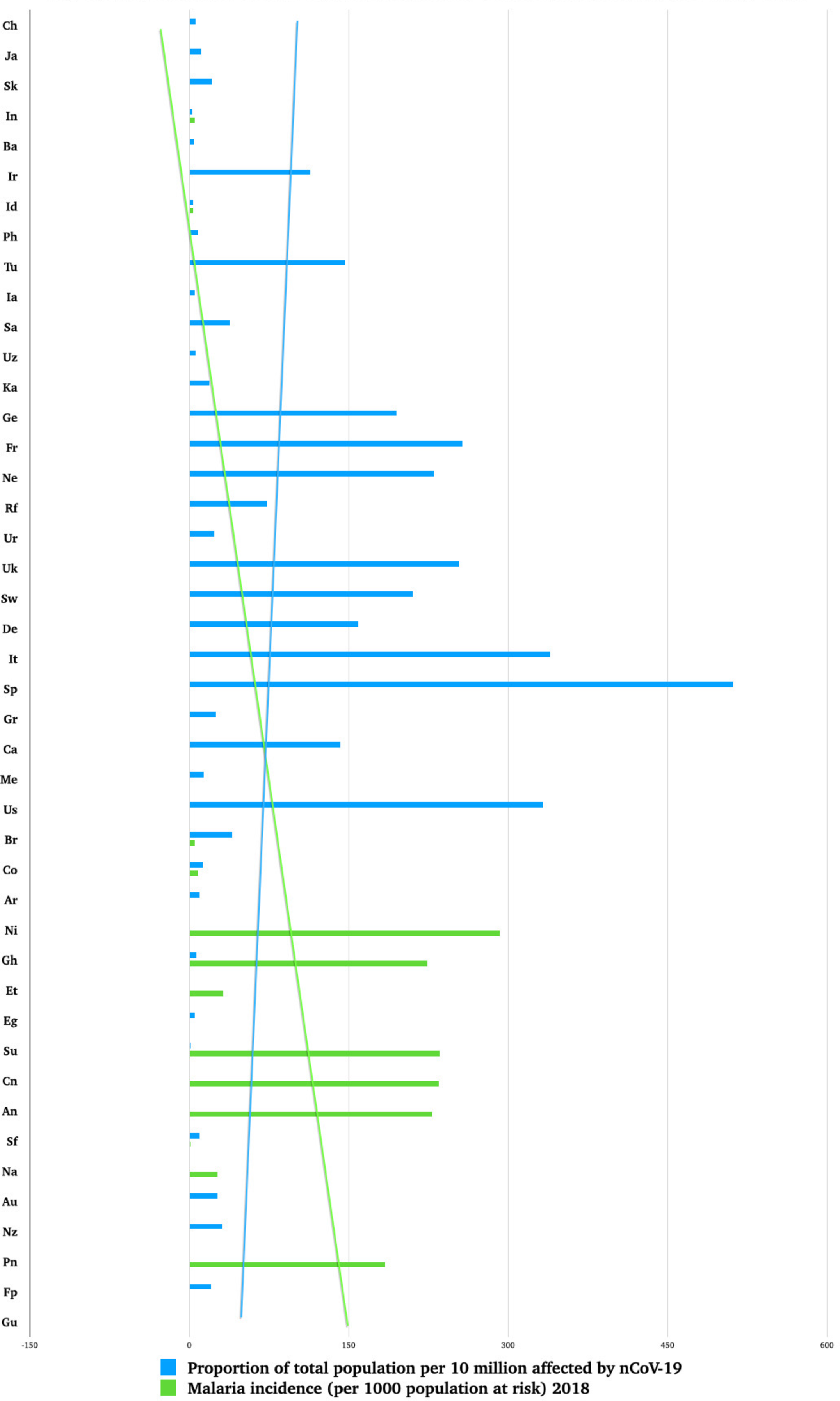
Proportion of total population affected vs Malaria incidence, country-wise.

The percentage of the population affected with Covid-19 decreases with higher incidence rates of tuberculosis (r =—0.4), as demonstrated in Figure 4. Also the height of the peak (the highest number of cases reported in a day) is observed less with increasing incidence of tuberculosis (r=—0.25).

**Fig 4.**
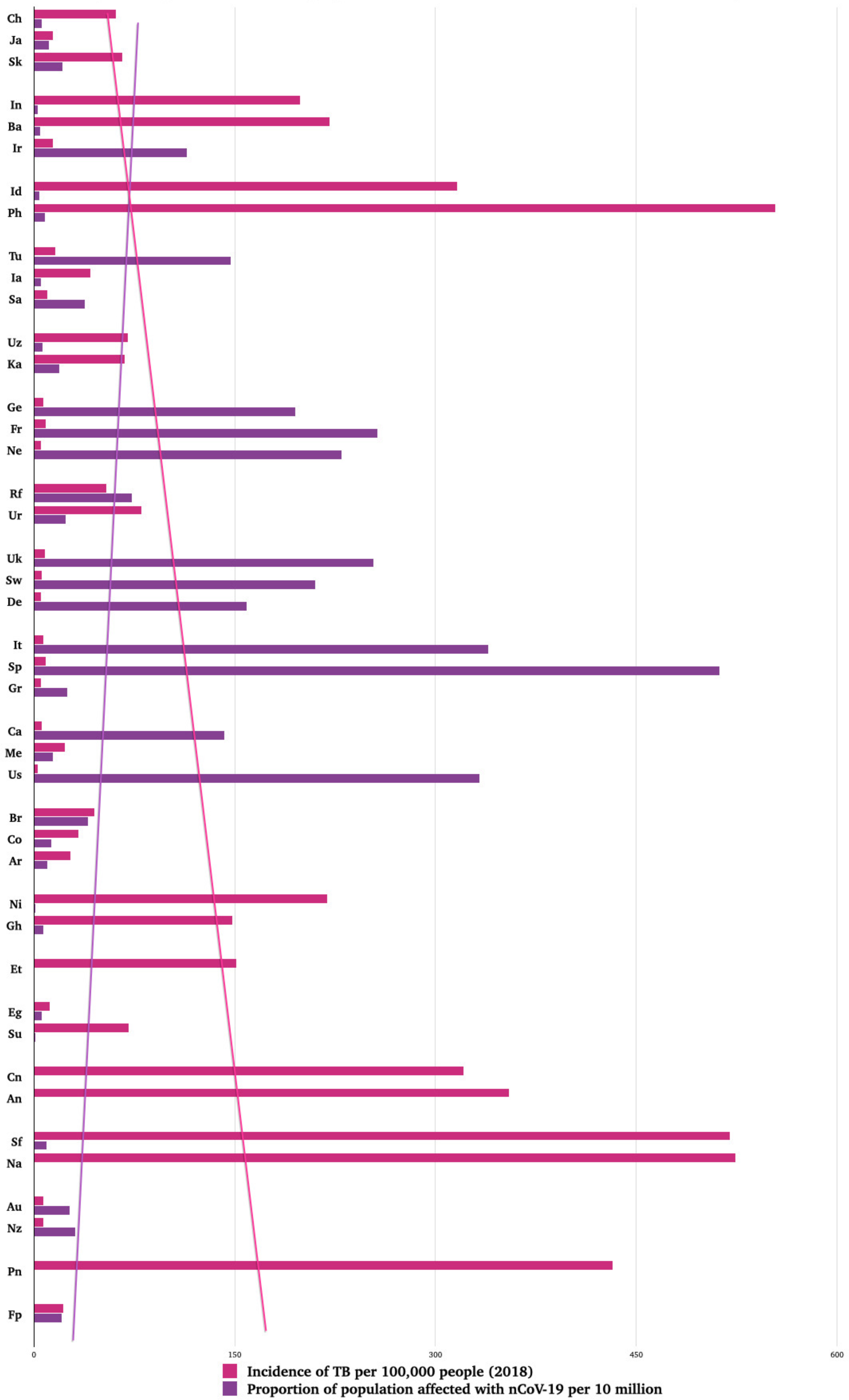
Proportion of total population affected vs incidence of TB, country-wise.

**Fig 5.**
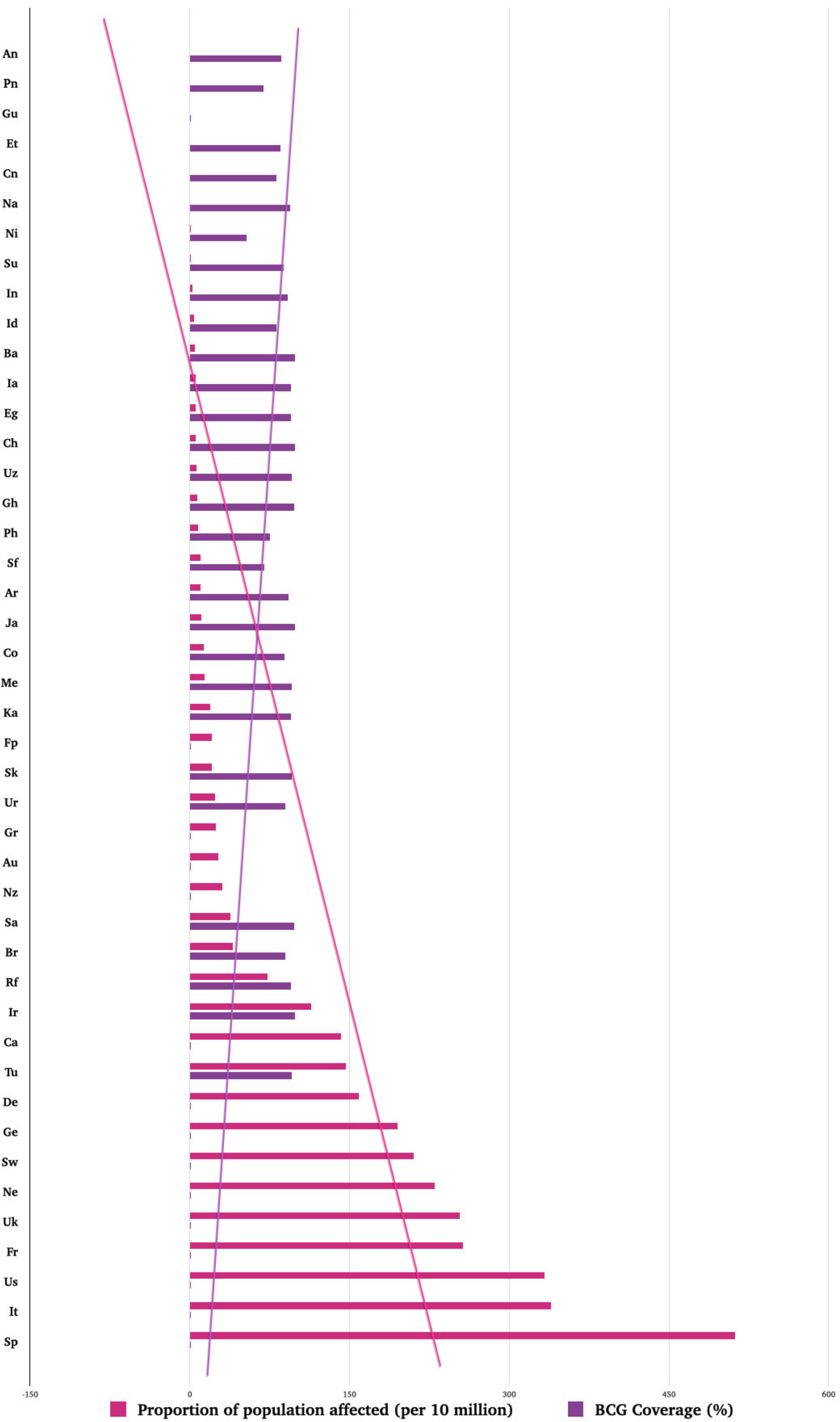
Proportion of population affected vs BCG coverage, country-wise.

The percentage of the population positive for Covid-19 varies inversely with the BCG coverage of the country (r=—0.63) as seen in Figure 5, assuming those with no universal BCG programme having 0.01% coverage). The total number of Covid-19 cases reported is decreases with higher BCG coverage (r=—0.33). The average Covid-19 case fatality rates are also lower (r=—0.24). Earlier introduction of BCG vaccination programmes are associated with lower total number of Covid-19 cases in countries with universal vaccination programmes.

The average number of days until the spike was 49 days for countries enforcing complete stringent lockdown, 61 days for those with selective stringent lockdown, 54 days with moderate and 70 days with nominal lockdown. The USA, was an outlier with 100 days till the spike following moderate lockdown measures. However the weighted average of the percentage of the population diagnosed with Covid-19 was higher in the moderate lockdown measures category (0.0026) compared to complete stringent lockdown (0.0003).

A more gradual curve (longer interval from second highest peak day till peak day) is weakly associated with a lower peak number of cases reported on peak day (r= —0.12). The more the number of days until the spike the higher it was (r=0.39), that is the number of cases reported on peak day was greater. The case fatality rate increased when the duration to the spike was prolonged (r=0.26).

## Discussion

The correlation between the percentage of cases in the total population and the median age of the population indicates that the diagnosis of the disease is more likely in older adults, which may be due to increased symptom burden and different care-seeking behaviour.

That there is no geographical correlation between population density and percentage of the population affected by Covid-19, shows that it is incorrect to assume that regions with high population densities would be incapable of abiding by social distancing norms and would thus inevitably report high incidence of the disease.

The finding that a higher percentage of population affected in certain climate zones was then corroborated by studying the climate characteristics of regions in the USA and India that were more severely affected than other parts of those countries. It was noted with interest that the New York state of the USA which is worst hit with the maximum percentage of the population affected (0.037%) is in the Aw, BSh climate zone, the same as Maharashtra, the most affected state in India. California (50,129 cases on 30/4/20) and Washington (14,327 cases on 30/4/20) also with a high number and proportion of cases of Covid 19 also belong to the same climate zones. The climate zones in Illinois (52918 cases on the 30/4/20) in Florida (33,690 cases on 30/4/20) are Aw Cwa while in Pennsylvania (47,999 cases on 30/4/20) they are Cwa, BSh. Delhi - the capital city of India, also Cwa, BSh, is experiencing a proportionately larger number of cases, as well as the Indian state of West Bengal (Aw, Cwa). Among the European countries Spain, France, the UK, Netherlands have Oceanic (Cfb) as the most prevalent climate type while Italy has Cfa (Humid subtropical), and while Cfb is the second most prevalent climate type in Germany and Denmark, the most prevalent is Dfb (Humid continental). It seems that the proportion of the population affected is comparatively less in Af (Tropical rainforest), Am (Tropical monsoon), BWh (Arid hot), Dfc (Sub-arctic) climate prevalent countries.

Widespread therapeutic and prophylactic use of chloroquine and hydroxychloroquine in areas of malaria endemicity, associated with higher physician comfort in instructing off-label use of these drugs for Covid-19 suspects and cases, may be causally associated with the reduced incidence of Covid-19, and subsequent lower case fatality rates in these countries. Prescription of antimalarial drugs on the basis of clinical suspicion for fevers of unknown origin where laboratory diagnosis is not immediately possible, is a common practice in malaria endemic regions, and affords good clinical outcomes.

The strength of the negative association between BCG coverage and country-wise case fatality rates of Covid-19 (r=-0.33) is almost equal to the correlation between median age and case fatality rates (r=0.32). The increased risk of mortality in older age groups has widely been accepted by the WHO and the scientific community. Hence, statistically the protective effect of BCG vaccination is as strong as the impact of morbidity enforced by increased age. The increased mortality with age might also be explained in part by the waning of vaccine efficacy of the BCG vaccine after 40 years^33^.

The finding of lower incidence of Covid-19 with BCG coverage and endemicity of malaria and TB might not be merely circumstantial evidence.When a virus infects cells of a body, the body immediately mounts a non-specific innate response consisting of neutrophils, macrophages and dendritic cells which slow the progress of the virus and may even prevent the development of symptoms. COVID-19^34^ may inhibit the expression of IFN-1 to overcome the immune response like its predecessor SARS-CoV Type I interferons are produced by immune cells in response to viruses and they act on infected as well as uninfected cells, activating enzymes that cause the degradation of viral nucleic acids and the inhibition of viral of replication, causing the induction of a kind of ‘antiviral state’^35^. Type 1 IFN is well known for its antiviral activity and stimulation of effector T cell responses ^36^. It has also been found that these Type 1 interferons are important regulators in developing anti-parasitic immunity against Plasmodium falciparum, the protozoal causative agent of malignant malaria^37^ ^38^.

It is also found that type 1 IFN signalling may contribute to the host defence against M. tuberculosis infection, though extensive type 1 IFN responses like in the context of viral infections could promote pathology of the disease^39^. There are reports of IFN-alfa boosting effects of BCG vaccinated mice^40^ and augmentation of BCG immunogenicity by IFN-beta (both subtypes of IFN type 1). Adaptive changes in IFN-1 expression in populations with Malaria and TB endemicity may lead to lower incidence of Covid-19. The administration of BCG can also change the cytokine profile in subjects leading to the reduced incidence of Covid-19 in regions reporting higher BCG coverage. The missing link in this may be recent RCTs^41^ demonstrating the induction of immunity and protective efficacy of the anti-tubercular vaccine BCG against controlled human malaria infection due to changes in the innate immune system that lead to earlier phenotypic monocyte activation and expression of NK cell activation markers that is inversely correlated with parasites on malaria challenge. A recently published multi centre prospective randomised trial^42^ in Hong Kong has found IFN beta-1b, a Type 1 IFN, to be effective in combination with the antiretroviral lopinavir-ritonavir and the antiviral ribavirin reducing the mean duration of disease (start of treatment till negative nasopharyngeal swab) lending further strength to the hypothesis that higher IFN type 1 levels would lead to lower manifestation of symptoms and reduced morbidity from the Covid-19 disease.

The choice of mitigation measures by different populations seem to be more of a response to the epidemiological curve rather than a determining factor. This explains why no clear trend of delayed and diminished peaks are seen with stricter social distancing norms. Data about lockdown measures has been collected as a measure of the respective country’s response to the onslaught of the epidemic. This data may be useful in later studies to compare the impact of the epidemic itself with the magnitude of the impact of our collective response.

A study of acute antibody responses to SARS-CoV-2^43^ in 285 patient found all patients tested positive for IgG within 19 days after symptom onset. However total measurable antibody is not the same as functional protective antibody. No study has concretely evaluated whether the presence of antibodies to SARS-CoV-2 confers immunity to subsequent infections by the virus^44^. On a community scale this means it cannot yet be assumed that if infection and subsequent immunity levels in populations reach the critical proportions needed for the achievement of herd immunity it would halt the epidemic. It seems prudent to add that if it is indeed found that the detected IgG do confer immunity to reinfection over a period of time (1-4 years based on studies of closely related coronaviruses) at least 60% of the population would need to have protective immunity (considering an R0 of 2.2, with estimates being higher if R0 is underestimated^45^, and varying with effective reproduction number or R_t_ from population to population) in order to halt further spread in that population^46^. The number of people who need to be vaccinated in order to attain herd immunity were also calculated based on an Ro of 2.2. The huge populations living in developing and underdeveloped countries will need to have access to the vaccine in sufficient numbers for any protective effect at the community level. This would need planning and forethought to ensure that once the vaccine is available it can be administered as soon and as widely as is necessary to truncate the effect of the epidemic. Having said that, it will be prudent to assume that to achieve the critical number through vaccination would require decades of immunisation effort. It can be summarised that even if there is this effective vaccine with no/less negative longterm or short-term health implication, there is no quick escape from this situation. Which strengthens the necessity of multiple reviews of our existing arsenal of mitigation of any health emergency rather that being consistently sceptical about those. It is evident that it is less likely to have one single and absolute solution to this pandemic, so it’ll be wise to look for a multi-pronged and collaborative assembly of probable solutions, which is the ultimate goal of this study.

### Limitations

We could not obtain excess mortality rates for the time period of the epidemic for many of the selected countries. Attributable risk from BCG vaccination could not be calculated because health data of the infected population could not be sourced in the public domain. We could not obtain complete data of therapeutics being used or of mitigation and alleged suppression tactics used in different countries. All the data was sourced from public domain websites which may have some inaccuracies of their own.

## Conclusions

in confirmation of trends suggested by empirical data this study found a strong geographical correlation between median age and total number of cases of Covid-19 and also a positive relationship between median age and the average death rate. Data from 45 countries spread over all inhabited continents were analysed to find that a larger proportion of cases were occurring in Oceanic (Cfb), Humid subtropical (Cwa, Cfa), Humid continental (Dfb), Tropical Savannah (Aw), and Semi Arid hot (BSh) climate zones. Confirming empirical data it was found that global trends indicated that higher malaria incidence correlated well with lower case burden of Covid-19. The prevalent use of chloroquine and hydroxychloroquine in malaria endemic zones could be a reason for the reduced case burden and burden of mortality from Covid-19 in these areas. The protective effect of the BCG vaccine was quantified in terms of its correlation with reduced incidence of Covid-19 and was found almost equal in magnitude to the deleterious effect of age. it has been hypothesised that in addition to trained immunity, higher levels of Type 1 interferon was responsible for this protective effect. More stringent social distancing measures were also found to have a positive effect on disease control.

However it is important to note that the countries of Africa, Asia and South America with lower median ages and higher incidence of malaria and tuberculosis, along with higher BCG coverage already have lower life expectancies, and strained health systems, with much lower health system performance indices which may eventually cancel out any protective effects of these combination of favourable factors in case of a typical massive outbreak. So while we cannot derive any reassurance of reduced impact in the poor and developing nations, as the threshold of number of total cases needed to surge might fall far too short of the need based projected and report based actual strength of their existing health systems. But this study holds that the discussed impact of outliers might as well be the reasons behind any reduction to the expected load of cases in those countries, rather that attributing those reductions unto ‘unknown’ factors. Given that possibility we can inclusively use this data and study to better understand the course the disease might follow in the future. The findings of the study also underline the need for more widespread clinical trials of hydroxychloroquine and the BCG vaccine in the prophylaxis and treatment of Covid-19. The rapid and long lasting immunological modifications caused by the BCG vaccine which confer some protection to viral respiratory tract infections may be put to use by administering BCG prophylactically to health care workers and high risk populations. The adverse effects of BCG vaccination are few and its long term effects are well documented. The side effect profile of chloroquine and hydroxychloroquine are also well known to the medical community and can be avoided by simple precautions before prescribing. On the other hand the cost of developing and producing a new vaccine in sufficient numbers needed for a herd immunity are significantly higher and the long term effects of any new vaccine would be unknown in spite of clinical trials. The human cost and suffering of continuing economic lockdown until the development of a vaccine are unsustainable in the developing and poorer nations and the long term effects of the poverty and undernutrition induced by these mitigation efforts may equal or likely surpass the burden of morbidity and mortality from the disease itself. The evidence found in this study suggests that it would be far more rational to thoroughly explore the scope of using the BCG vaccine, chloroquine and hydroxychloroquine to reduce the incidence and morbidity of the disease in the population and thus spare the already strained capacities of health systems in the developing and poorer countries to whatever extent is possible.

## Data Availability

with publication

## Appendix 1: Master table of data collected

Refer Supplement

